# Genomic Evidence Links Inflammation to Residual Pulmonary Vascular Obstruction and Risk of Pulmonary Embolism Recurrence

**DOI:** 10.64898/2026.06.26.26356642

**Authors:** Floriane Samaria, Gaëlle Munsch, Ohanna C. L. Bezerra, Kerri L Wiggins, Lénaïck Gourhant, Astrid van Hylckama Vlieg, Marine Germain, Robert Olaso, Ilana Caro, Noémie Saut, Delphine Bacq, Catherine A Lemarié, Stéphanie Debette, Nicholas L Smith, Frits. R Rosendaal, Pierre-Emmanuel Morange, Grégoire Le Gal, Jean-François Deleuze, France Gagnon, Marc A Rodger, Francis Couturaud, David-Alexandre Trégouët

## Abstract

**Background and Aims:** Residual pulmonary vascular obstruction (RPVO) defined as the persistence of thrombotic material within the pulmonary arteries several months after an acute pulmonary embolism (PE) is associated with an increased risk of severe complications, including recurrent events and chronic pulmonary hypertension. However, the genomic architecture underlying RPVO in unprovoked PE remains poorly understood, and this study aims to address this gap.

**Method:** By leveraging genetic and imaging RPVO data from three independent cohorts totaling 586 unprovoked PE patients, we conducted a meta-analysis of genome wide association study (GWAS) of RPVO using a dedicated statistical method to handle the semi-continuous distribution of RPVO. The meta-GWAS was complemented by haplotype association analyses and transcriptome wide association studies as well as Mendelian Randomization (MR) approaches based on plasma metabolites and proteins.

**Results:** Through meta-GWAS, we identified one locus, *OSTN*, associated with RPVO (lead variant rs59109356 associated with a ∼2-fold increase of RPVO, p=3.92×10^-8^). A second locus, *CCN4*, previously reported to associate with pulmonary fibrosis, was also identified, with evidence of association approaching genome-wide significance (p=6.7×10⁻⁸). We also identified a common haplotype spanning over *AHSG/HRG/KNG1* associated with a ∼3-fold increase of RPVO (p=2.96×10^-8^). Using plasma protein-based MR, we demonstrated that one unit increase in genetically determined plasma levels of IL-1 R AcP encoding IL1RAP was associated with a 28% (p=1.32×10^-6^) reduction in RPVO. We also observed statistical evidence that the *CCN4* (p=0.06) and *IL1RAP* (p=0.02) loci associate with the risk of PE recurrence in a sample of 1,617 unprovoked PE patients.

**Conclusionsa:** By identifying novel molecular determinants of RPVO that map to loci involved in inflammatory pathways and vascular remodeling, our study provides evidence that inflammation is the predominant—and likely the key—mechanism underlying RPVO, whereas impaired fibrinolysis appears to play a more limited role.

## Introduction

Over the last decades, the management of pulmonary embolism (PE) has mainly focused on the acute event, including its diagnosis, patient care and short-term prognosis (1). This focus is driven by the increasing incidence of PE, its status as the third most common cardiovascular diseases worldwide and the high risk of serious or fatal complications occurring in the days following the event (2–8). Nevertheless, PE survivors may suffer from long-term complications, more particularly patients with unprovoked PE, defined as PE occurring without any identified major risk factors (9–11). Following their 3 to 6 months initial anticoagulant therapy, up to 30% of these patients have an incomplete resolution of thrombus resulting in a persistent residual pulmonary vascular obstruction (RPVO) (12–18). This suggests that the remaining clot is transforming into permanent fibrous scar tissue and that this occurs probably early after the acute event (19). It has already been shown that RPVO is a strong predictor of recurrence of PE, chronic thromboembolic hypertension (CTEPH), right ventricular dysfunction and other cardiac impairment (1,14,17,19–27).

The mechanisms underlying the remodeling of an acute thrombus into permanent fibrotic scars remain poorly understood, although hypotheses have been proposed. Some studies suggested that impaired fibrinolysis may play a central role in scar formation through reduced fibrin degradation and dysregulated expression of key fibrinolytic components (19,22,28–31). Similarly, Planquette et al. proposed that RPVO is likely related to incomplete thrombus resolution, analogous to the residual vein obstruction observed after deep vein thrombosis (17). They also emphasized that persistent perfusion defects have not been associated with a procoagulant state; rather, they may reflect resistance to fibrinolysis in patients with PE (17,32). More recently, Shah K. P. et al. suggested that the inflammatory response occurring during the acute thrombotic episode may induce permanent pulmonary arterial damage, leading to vascular wall thickening and fibrotic remodeling (20). Finally, several hemostatic and fibrin-related biomarkers, including fibrinogen β-chain monosialylation, fibrin clot permeability, endogenous thrombin potential, clot lysis time and fibrin clot permeability have been associated with RPVO and proposed as potential contributors to its development (33).

To date, only a few relevant risk factors have been identified as promoting RPVO, such as advanced age (11,15,16,34), unprovoked PE (16,34), elevated initial pulmonary vascular obstruction at PE diagnosis (11,15,34), chronic respiratory disease (34), prior venous thromboembolism (VTE) (11) and elevated factor VIII levels (34). While genetic susceptibility has been demonstrated for many of these risk factors (35,36), as well as for RPVO-related complications (37,38), the genetic determinants of RPVO itself have not been systematically investigated. To address this gap and improve our understanding of the biological mechanisms underlying RPVO, we here present the results of the first genome-wide association study (GWAS) on RPVO. This analysis was complemented by downstream genomics-driven approaches, leveraging GWAS data together with RPVO imaging data from three independent clinical cohorts.

## Methods

### Studies and participants

To investigate the genetic basis of RPVO, we implemented a GWAS meta-analysis framework leveraging genetic and RPVO imaging data from three clinical cohorts: the French EDITH (*Etude des Déterminants et Interactions de la THrombose veineuse)* cohort (15,41), the French randomized clinical trial PADIS-PE (Prolonged Anticoagulation During eighteen months vs placebo after Initial Six-month treatment for a first episode of idiopathic Pulmonary Embolism) trial (NCT00740883) (34,42) and the Canadian cohort REVERSE-I (REcurrent VEnous thromboembolism Risk Stratification Evaluation) (43,44). For this work, we selected patients aged 18 years and older who had a first episode of unprovoked-PE, completed their initial 3-6 months anticoagulant therapy, underwent a ventilation/perfusion (V/Q) scan, and were genotyped for genome-wide common polymorphisms as recently described (45). Following quality control of the genetic data (45), the final sample sizes were 306, 165, and 115 patients for PADIS-PE, EDITH, and REVERSE-I, respectively.

Unprovoked PE was defined consistently across the three studies as the absence of cancer within the 12 months preceding the PE event and of lower-limb trauma, prolonged immobilization ≥72 hours for an acute medical illness, surgery with general anesthesia ≥30 minutes, pregnancy, post-partum status or estrogen-containing pill within the past 3 months of PE.

RPVO, the primary outcome of the present GWAS, was assessed using V/Q scan following a previously detailed protocol (46) and expressed as percentage of global pulmonary arterial obstruction. In the REVERSE-I and PADIS-PE studies, RPVO was assessed after completion of the initial 6-month anticoagulant therapy for PE. In contrast, in the EDITH study, RPVO measurement was performed between 3 and 24 months after the PE event. As shown in **Supplementary Figure 1**, the distribution of RPVO in each study is semi-continuous, characterized by zero inflation, with the remaining observations forming a positive continuous distribution extending up to 96%.

### GWAS and fixed effect meta-analysis

Association of single nucleotide polymorphisms (SNPs) with RPVO was tested using a Compound Poisson Gamma (CPG) model to handle the semi-continuous nature of the RPVO variable (45,47). In each study, association analyses were conducted under the assumption of additive allele effects and were adjusted for age, sex and the first principal components capturing more than 80% of the variance in genome wide SNP data. Only bi-allelic autosomal SNPs with a minor allele frequency (MAF) >1% and an imputation quality score greater than 0.3 were considered. GWAS analyses were conducted using R 4.1.0 software (R Core Team (2021), https://www.R-project.org/), with *cplm* 0.7.12 package (48) for CPG modelling. Manhattan and Quantile-Quantile plots were generated using the *qqman* and *EnvStats* R package, respectively.

For each SNP, regression coefficients estimated using the CPG model in the three studies were then combined through meta-analysis using the inverse-variance method as implemented in the GWAMA software (49). Exponentiated coefficients represent the multiplicative change in RPVO associated with the tested allele. Heterogeneity across studies was estimated using the I^2^ statistic (50). Only SNPs available in the three studies were retained for meta-analysis. A statistical threshold of p<5×10^-8^ was used to declare genome-wide significance.

To detect the presence of additional independent RPVO-associated SNPs at the identified genome-wide significant locus, we conducted, in each of the three studies, a conditional GWAS restricted to the corresponding chromosome. These analyses were adjusted for the imputed allelic dosage of the lead SNP to assess the presence of secondary independent association signals. We then meta-analyzed results following the aforementioned methodology used for the original GWAS.

### Transcriptome wide association study

Using the summary statistics of our RPVO GWAS meta-analysis, we performed a transcriptome-wide association study (TWAS) following the FUSION pipeline (51) from which only results from LASSO prediction models (52) were considered. Genetically determined gene expressions across 18 tissues available on the GTEx database (**Supplementary Table 1**) were analyzed. TWAS was complemented with conditional and colocalization analyses as implemented in FUSION. Associations of gene expressions with RPVO were considered significant if they reached a Bonferroni-corrected threshold of p<8.34×10^-6^, corresponding to the 0.05 type I error corrected for the average number of genes tested per tissue/cell types (N=5,995). We then prioritized results with a posterior probability of a shared causal variant (PP4) greater than 0.75.

### Mendelian Randomization

Using our RPVO GWAS summary statistics, we performed a series of Two-Sample Mendelian Randomization (MR) analyses (53). The MR methodologies included Inverse Variance Weighted (IVW) (54), Weighted Median (55), Egger (56), and MR-PRESSO (57) implemented in TwoSampleMR package (58) in R 4.5.2 software and MR-PRESSO R package (57).

*Plasma protein based MR* - To identify plasma proteins whose genetic determinants may be causally linked to RPVO, we leveraged proteogenomics GWAS results from the deCODE project (N=35,559) (59) and the Fenland study (N=10,708) (60), where respectively 4,907 and 4,979 proteins were measured with Somalogic platform. Also, we made use of 2,940 proteins measured in the UK Biobank study (N=34,557) (61) with the antibody-based Olink Explore 3072 proximity extension assay. In each study, we selected independent genome-wide significant (p<5×10^-8^) genetic instruments (i.e. protein quantitative trait loci -pQTL) using clumping for linkage desequilibrium (LD), with a window of 10Mb and r^2^=0.01 based on European reference panels (62). Only proteins with >2 instrumental variables were selected to compute MR estimates using the IVW method. Cis and trans-pQTLs were included as instrumental variables in these analyses.

*Plasma metabolites based MR* – A similar MR workflow was deployed on 1,091 blood metabolites and 309 metabolite ratios measured in N=8,299 unrelated individuals from the Canadian Longitudinal Study of Aging (63). After our selection strategy for genetic instruments (see above), 423 metabolites were selected for MR analysis in relation to RPVO. A Bonferroni-corrected threshold of p<1.18×10^-4^ was then used.

*MR on hemostatic traits* – We leveraged GWAS summary statistics on 30 hemostatic traits selected for their association with thrombotic phenotypes (**Supplementary Table 2**) that were available in GWAS catalog (https://www.ebi.ac.uk/gwas/downloads/summary-statistics) to conduct MR with RPVO following the same analysis plan as above. A Bonferroni-corrected threshold of p<1.7×10^-3^ (∼0.05/30) was used here to declare statistical significance.

### Correlation analysis between candidate plasma proteins

To estimate the genetic correlation between plasma proteins identified as associated with RPVO in our GWAS framework, we used the Linkage Desequilibrium Score regression (64,65) approach, as implemented in the LDSC software, using known LD scores for European ancestry individuals from 1,000 Genomes Project Phase 3 (https://alkesgroup.broadinstitute.org/LDSCORE/) (62). The same publicly available proteogenomics resources employed in our MR analysis were used. We also used individual plasma proteomic and GWAS data of an independent sample of 966 population-based participants of the 3C-Dijon study (66,67) profiled on the Olink Explore 3072 panel. These data were used to estimate the biological correlations between selected candidate proteins and to assess the impact of molecular markers associated with RPVO on plasma protein levels.

### Candidate gene analyses

In addition to the aforementioned MR analysis, we conducted analyses to assess whether genetic variants at well-known genes involved in coagulation and fibrinolysis pathways (68,69) (**Supplementary Table 3**) might be individually associated with RPVO. For each gene, we defined a corresponding genomic locus based on gene annotations, extending ±500Kb around the gene to capture potential cis-regulatory variants.

We also assessed whether genetic variants identified in GWAS for VTE (70), CTEPH (40) and PE recurrence (39) were also associated with RPVO.

### Impact of genomic findings on PE recurrence and CTEPH

The molecular determinants we identified for RPVO were subsequently evaluated for their association with recurrence of unprovoked PE and the risk of CTEPH.

For PE recurrence, we leveraged individual genetic data of unprovoked PE patients from 6 studies including the 3 studies used for the RPVO GWAS plus the Marseille Thrombosis Association (MARTHA) (71), the Multiple Environmental and Genetic Assessment (MEGA) (72), and the Heart and Vascular Health case-control study of VTE (HVH) (73). For CTEPH, we used summary statistics from the recently published GWAS on CTEPH (40).

In addition to testing the impact of SNPs on PE complications, we also investigated the impact of the proteins identified in our MR analysis. For this, we further conducted a meta-analysis of the summary GWAS statistics obtained in UK Biobank, deCODE and Fenland. Using the same clumping based strategy as that mentioned above, a weighted Polygenic Risk Score (PRS) expected to serve as a surrogate marker for genetically determined plasma levels of our protein of interest. This PRS was then tested for association with recurrence in unprovoked PE patients using Cox model analysis (39).

## Results

A brief description of the characteristics of the 586 unprovoked patients with RPVO and genetic data used in this meta-GWAS analysis is available in **Supplementary Table 4**.

### GWAS fixed-effect meta-analysis on RPVO

After quality controls, 10,335,565 SNPs were included into the meta-analysis for association with RPVO. The results summarized in the Manhattan Plot (**Figure 1**) highlight a single genome-wide significant locus on 3q28. The Quantile–Quantile plot did not indicate any notable inflation (**Supplementary Figure 2**), with a genomic inflation factor of λ=0.99. Full metaGWAS results are available on the GWAS catalog while all association results with p<5×10^-6^ are given in **Supplementary Table 5**.

**Figure 1.**
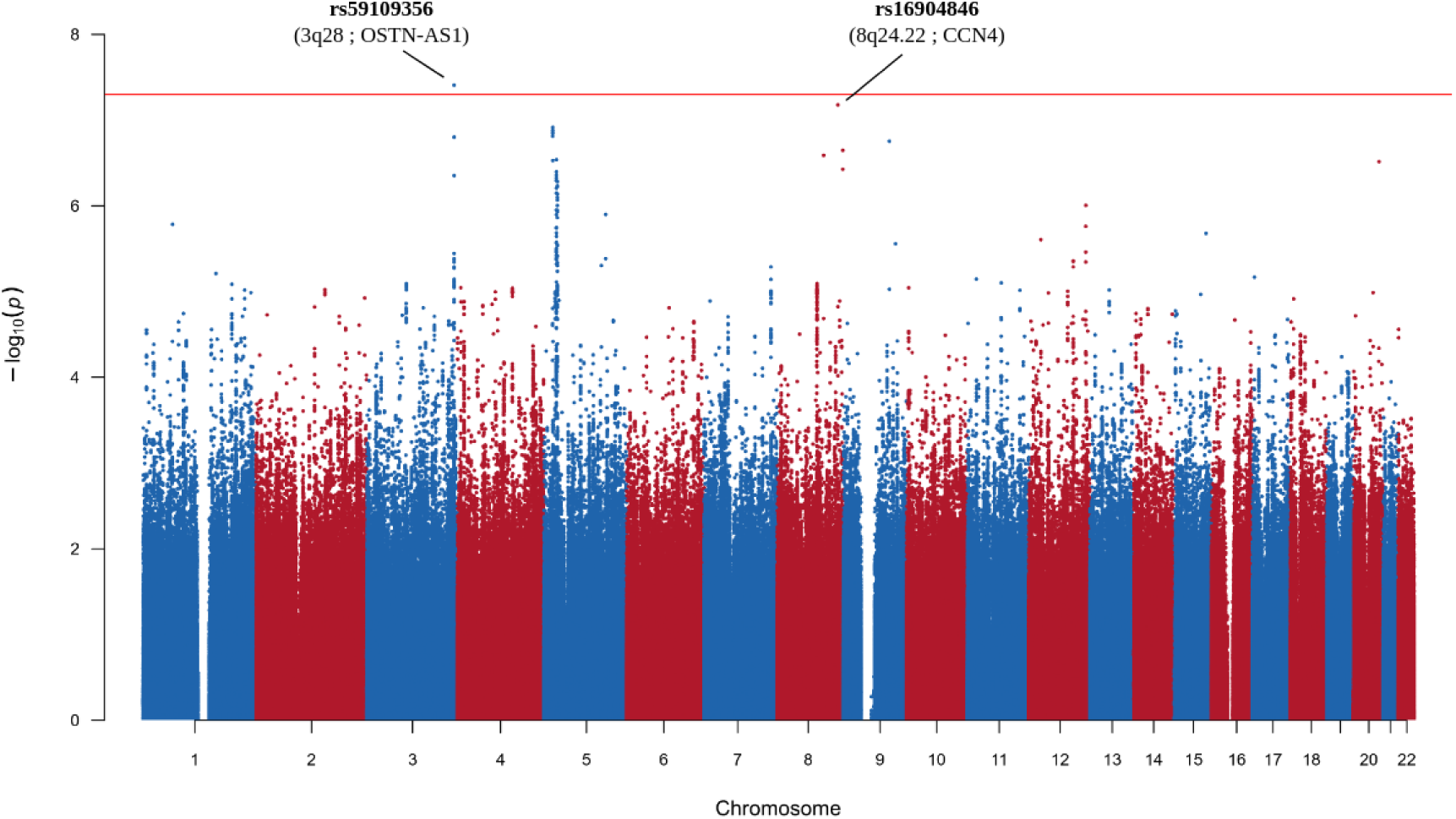
Manhattan plot of the GWAS meta-analysis results on RPVO (N=586). The horizontal red line represents the genome-wide threshold (P<5×10^-8^). Two main loci of interest located on chromosomes 3 and 8 are annotated.

The lead SNP on chr3q28 locus is rs59109356, located in intron 2 of *OSTN.* The minor C allele (frequency ∼0.11) was associated with a 1.98-fold increase of RPVO [95% Confidence Interval: 1.56 – 2.49] (p=3.92×10^-8^) (**Figure 2**). Allele effect estimates were consistent across the three contributing studies. As shown in **Supplementary Table 6**, consistent with the observed association, carriers of the rs59109356-C allele were more likely to exhibit RPVO and showed higher RPVO (>0) levels than non-carriers.

**Figure 2.**
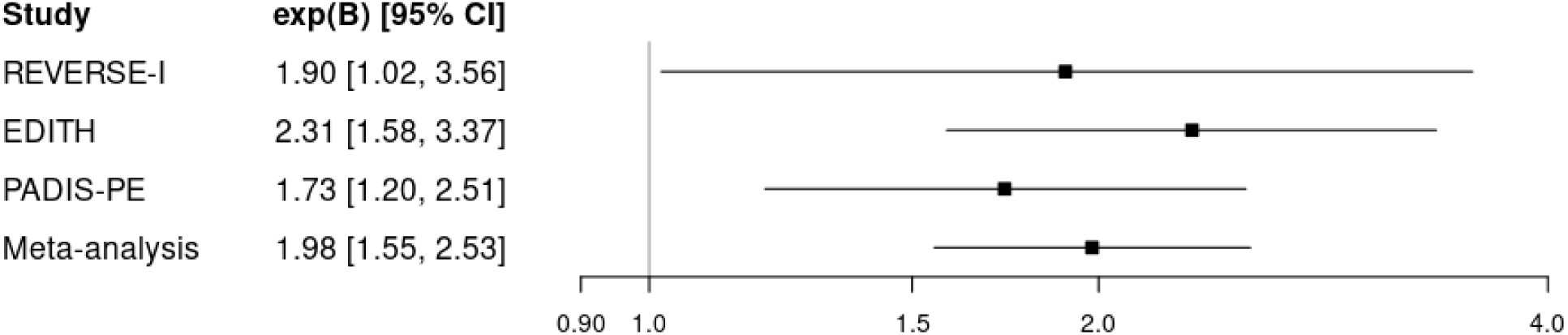
Forest plot of rs59109356 associations* with RPVO in EDITH, PADIS-PE, REVERSE-I and meta-analysis. I2=0; Heterogeneity p-value=0.56; *Associations were tested using a compound poisson-gamma model adjusted on age, sex and the four first principal components, under the assumption of additive allele effect.

After conditioning on the rs59109356-C allele, the smallest p-value observed at the *OSTN* locus (± 500kb) was p=2.65×10^-3^ **(Supplementary Table 7),** suggesting that the statistical signal originally observed was likely driven by a single SNP effect. The corresponding regional association plot at the *OSTN* is shown in **Supplementary Figure 3**. According to the GTEx portal, the C allele shows statistical evidence (p<10^-4^) for association with increased *OSTN* expression in different tissues (i.e. subcutaneous adipose, skeletal muscle and sun exposed skin). By leveraging plasma proteogenomics results from Fenland, DeCODE and UK Biobank, we observed a significant (p=4.27×10^-7^), albeit heterogeneous, cis effect of rs59109356 on plasma levels of Osteocrin, the protein encoded by *OSTN*. Plasma levels of no other proteins were found to be influenced by rs59109356 (p > 0.04 for all proteins except in the combined meta-analysis) (**Supplementary Table 8**).

A meta-analysis of Osteocrin GWAS summary statistics across Fenland, DeCODE and UK Biobank, identified the rs13322016 as the strongest cis pQTL for Osteocrin (p=2.37×10^-22^). This polymorphism showed a modest statistical association with RPVO (p=3.57×10^-3^) that vanished (p=0.290) after conditioning on rs59109356 (**Supplementary Table 9**).

Of note, the second most significant locus which narrowly failed to reach genome-wide significance (p=6.7×10^-8^) in the meta GWAS analysis maps to *CCN4* on 8q24.22 (**Supplementary Table 5**). The lead SNP is the uncommon intronic rs16904846 where the C allele (frequency ∼1%) is associated with a ∼4-fold increased values of RPVO. As shown in **Supplementary Table 10**, 90% (=17/19) of the carriers of the rs16904846-C allele have positive RPVO value and in patients with RPVO>0 the C allele was associated with increased RPVO values. Also known as *WISP1*, *CCN4* encodes a protein shown to be involved in pulmonary fibrosis (74,75), making this locus a strong candidate to be further explored. Although rs16904846 is not reported in GTEx to associate with any gene expression levels in any tissue, it is found to be associated with *WISP1/CCN4* plasma protein levels in deCODE (β=0.27 [0.19-0.35], p=5.45×10^-11^) and UK Biobank (β=0.22 [0.16-0.27], p=1.32×10^-14^) studies.

### Downstream TWAS and MR analyses

Neither the multi-tissue TWAS analyses, whose results are presented in **Supplementary Table 11**, nor the MR analyses on metabolites **(Supplementary Table 12**) and on hemostatic traits (**Supplementary Table 13**) identified any new molecular marker (transcripts, metabolites, biological traits) associated with RPVO. Moreover, TWAS provided only marginal statistical support for an association of inferred *OSTN* expressions with RPVO (minimal p_TWAS_=8.18×10^-4^, pp3=0.65, pp4=0.29).

By contrast, MR analyses on plasma proteins revealed one novel molecular determinant of RPVO (**Supplementary Table 14**). Indeed, higher genetically predicted plasma levels of IL-1 receptor accessory protein (IL-1R AcP) were consistently associated with lower RPVO across the UK Biobank, Fenland, and deCODE proteogenomic resources. In UK Biobank, one increase unit of genetically determined plasma levels of IL-1 R AcP was significantly associated with a 34% decreased RPVO (exp(β)=0.66 [0.55 – 0.78], p=1.45×10^-6^). The same pattern was observed in Fenland (exp(β)=0.65 [0.54 – 0.77], p=1.36×10^-6^) and in deCODE (exp(β)=0.77 [0.66 – 0.89], p=5.91×10^-4^]). Meta-analysis of IL-R AcP GWAS summary statistics across the three resources **(Supplementary Table 15)** further strengthened this association, with a one unit increase in genetically determined plasma levels of IL-1 R AcP associated with a 28% (exp(β)=0.72 [0.63 – 0.82], p=1.32×10^-6^) reduction in RPVO.

Most of the genetic instruments of IL-1 R AcP (45 out of 65) are cis pQTLs mapping to *IL1RAP* on 3q28, approximately 600kb from the *OSTN* locus identified in the GWAS analysis (**Supplementary Table 15**). The lead variant, rs59109356, showed no LD with IL-1 R AcP independent pQTLs (**Supplementary Table 16**). Furthermore, joint analysis of rs59109356 with PRS derived from 65 pQTLs for IL-1 R AcP plasma levels confirmed the independent additive effects of these two loci (**Supplementary Table 17**). In the joint model, the rs59109356-C allele remained associated with 1.93-fold increased (95% confidence interval [1.52 – 2.44], p=4.5×10^-8^) of RPVO values and one standard deviation increase in a PRS derived from the 65 IL-1 R AcP genetic instruments was associated with 0.78-fold decreased of RPVO ([0.69 – 0.88], p=6.7×10^-5^) (**Supplementary Table 17**). The correlation between genetically determined OSTN and IL1RAP plasma protein levels estimated from UK Biobank was very low (r=0.03, p=0.96). Correlation estimates were not obtained from deCODE or Fenland owing to the low heritability of OSTN plasma levels. To further assess the relationship of these two proteins, we used a sample of 966 healthy participants of the 3C Study (66,67) in which OSTN and IL1RAP protein levels were quantified using the Olink technology. After adjusting for age and sex, the biological correlation was very modest (r_g_=0.07, p=0.020). Altogether, these results support independent contributions of the OSTN and IL1RAP loci to RPVO.

To follow up on the single effect observed at *CCN4* rs16904846, we also meta-analyzed the GWAS summary statistics for CCN4 plasma levels as done for IL1RAP. The resulting MR **(Supplementary Table 18)** did not show any evidence for a possible causal link between CCN4 plasma levels and RPVO (p=0.64). Note, as shown in **Supplementary Table 9**, the very low number of genetic instruments (n=2) for Osteocrin plasma levels prevented us from performing a reliable MR between osteocrin and the risk of RVPO.

### Candidate gene analysis

5 of 44 coagulation/fibrinolysis loci demonstrated suggestive statistical (p<10^-3^) and homogeneous associations with RPVO (**Supplementary Table 19**). Two pertained to platelet genes, *GP1BA* (p=6.8×10^-6^) and *GP5* (p=3.2×10^-4^) genes. A third one was observed with the missense (p.Glu1429Asp) rs9534264 mapping to *ZC3H13*. The rs9534264-T allele that associates (p=3.37×10^-4^) with a 1.43-fold increase of RPVO was significantly (p<10^-300^) associated in UK Biobank with increased plasma levels of Thrombin-Activatable Fibrinolysis Inhibitor (TAFI) encoded by the nearby *CPB2* locus. Of note, rs9534264 is in strong LD (r^2^=0.90) with the *CPB2* rs1926447 (Ile347Thr) known to regulate TAFI levels and reported to be associated with thromboembolic diseases (76). An MR-based analysis using UK Biobank proteogenomics data suggests a possible causal association between increased TAFI levels and RPVO (MR regression coefficients β=0.40 ± 0.13, p=0.002). However, no such trend was observed using deCODE (β=-0.05 ± 0.15, p=0.75) nor Fenland (β=0.05 ± 0.12, p=0.67) results.

The remaining two suggestive associations mapped to 3q27.3, nearby *KNG1,* (p=4.79×10^-5^) and to C1R/C1S (p=2.78×10^-5^). For these two loci, two independent SNPs were identified by the pairwise r^2^-based clumping approach. Haplotype analyses at these two loci (**Supplementary Table 20**) were further conducted. While these analyses confirmed the possible independent effects of the two SNPs (rs75767107, rs117827303) at the *C1R/C1S* locus, these analyses demonstrated that data at 3q27.3 were more compatible with the effect of a specific haplotype. Indeed, the haplotype composed of both rare alleles at rs58550067 and rs62292610 was associated with a genome-wide significant 2.72-fold ([1.92 – 3.85], p=2.96×10^-8^) increase of RPVO compared to the haplotype composed of the common alleles at both SNPs. These two variants are located on either side of the *KNG1* gene and, according to HaploReg online tool (77), do not exhibit strong LD with *KNG1* variants. *KNG1* is a well-known locus in the regulation of plasma coagulation Factor XI (FXI), but identified haplotypes did not show any evidence for association with FXI plasma levels in the 3C study (p=0.723). We used proteomic and genetic data available in the 3C study to look for association of rs58550067/rs62292610 haplotypes with plasma protein levels to help fine-map the locus potentially driving the association with RPVO. At the pre-fixed p=1.7×10^-5^ threshold corrected for the number of tested proteins, only one significant association (p=2.4×10^-6^) was observed, and it was a trans pQTL haplotype effect observed with plasma levels of Scribble Planar Cell Polarity Protein (**Supplementary Table 21**).

None of the SNPs previously reported in GWAS analyses to associate with incident (**Supplementary Table 22**) or recurrent VTE (**Supplementary Table 23**) showed suggestive statistical evidence for association with RPVO.

### Impact of genomic findings on CTEPH and PE recurrence

We further assessed whether the molecular determinants of RPVO identified through our GWAS framework associate with the risk of recurrence in unprovoked PE patients and with the risk of CTEPH.

As summarized in **Figure 3** and detailed in **Supplementary Table 24**, the *CCN4* rs16904846-C allele that associates with increased RPVO was borderline significantly (p=0.06) associated with a Hazard Ratio (HR) of ∼1.8 to develop recurrence. No association with recurrence was observed with *OSTN* rs59109356 nor with 3q27.3 haplotypes (**Supplementary Table 24**). For *IL1RAP*, a 1 SD increase in the PRS associated with decreased RPVO was linked to a significantly lower risk of recurrence (HR=0.88, p=0.02) (**Supplementary Table 24**).

**Figure 3.**
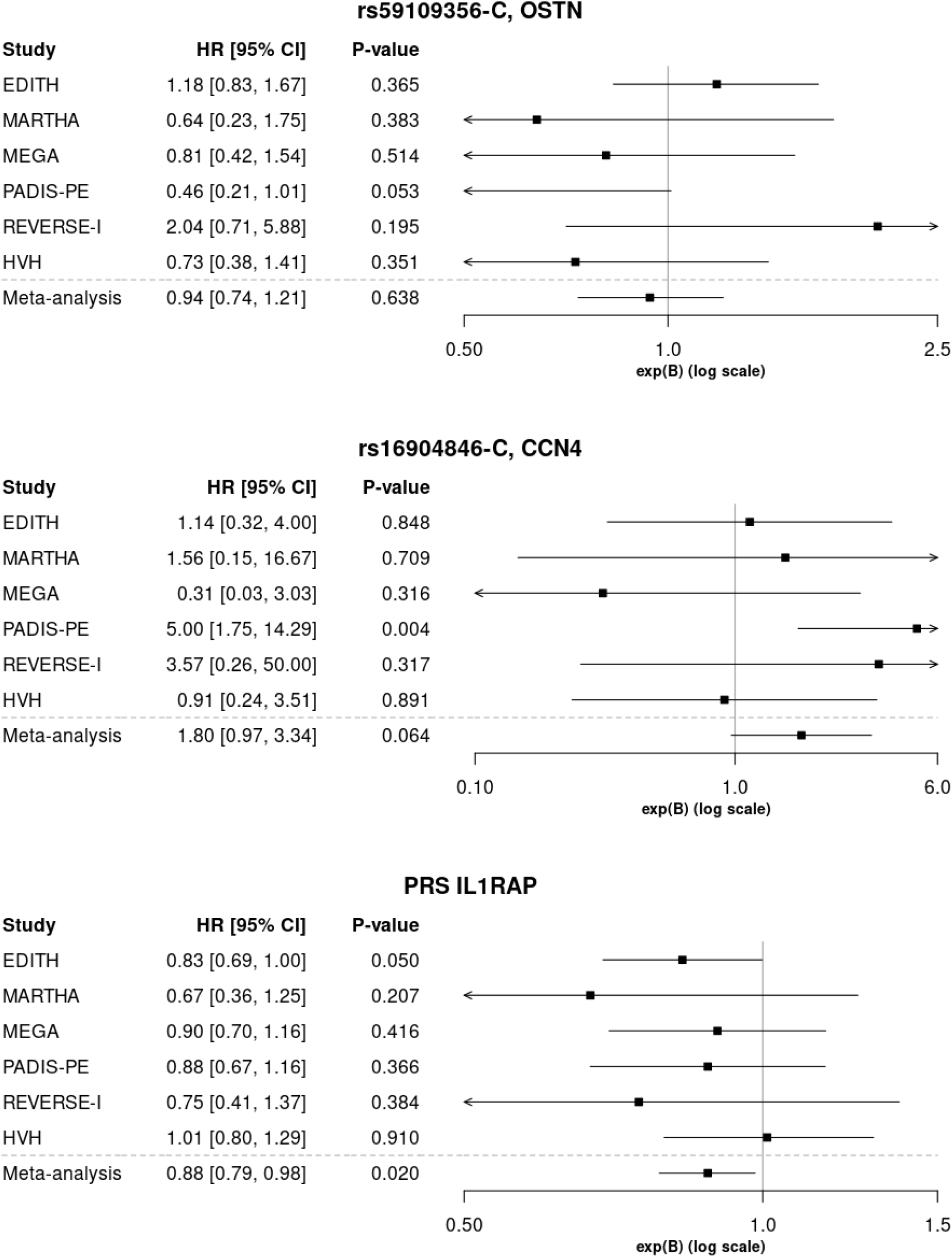
Associations* of rs59109356 (OSTN), rs16904846 (CCN4) and PRS IL1RAP with VTE recurrence (N=1,269, incl. 331 events). *Associations were estimated with a Cox model for VTE recurrence, adjusting for age, sex and the first principal components derived from the population stratification.

Regarding CTEPH, neither *OSTN* nor *CCN4* lead variants, nor the two variants rs58550067 and rs62292610 at the chr3q27.3 locus, were imputed in the publicly available GWAS summary statistics (40), preventing us from assessing their impact on the risk of CTEPH. Using CTEPH GWAS summary together with summary data from the meta-GWAS on plasma IL-1 R AcP levels in a MR framework, we found no evidence supporting an association between IL-1 R AcP plasma levels and CTEPH (β=0.002 ± 0.007, p=0.73).

## Discussion

By integrating GWAS, transcriptomic, proteogenomic, and MR approaches across three independent cohorts, we identified three significant molecular markers, *OSTN*, *IL1RAP* and *AHSG/KNG1*, both mapping to chromosome 3 (3q28 for the first two and 3q27.3 for the last one), associated with RPVO. A fourth borderline locus with strong biological support for its association with RPVO was identified, CCN4, on chr8q24.22.

*OSTN* encode Osteocrine, a structurally related to natriuretic peptides and may therefore interact with the natriuretic peptide system. This system contributes to the regulation of cardiovascular homeostasis (78,79) through modulation of cyclic guanosine monophosphate signaling, a key mediator against vascular remodeling (79,80). Osteocrine and natriuretic peptides have anti-fibrotic and metabolic effects by modulating inflammatory cytokines (78,81). In mice, osteocrine injection increased circulating plasma atrial natriuretic peptides levels, resulting in a reduced risk of cardiac remodeling after myocardial infarction (78,82). Similar effects on fibrosis and cardiac dysfunction have been observed in other experimental mouse models following injection of osteocrine (78). In other words, osteocrine is a protein that protects the vascular system by promoting the concentration of natriuretic peptides, which are the body’s natural defenses against fibrosis and heart failure. It is therefore reasonable to hypothesize that this protein could also contribute to clot remodeling following PE. However, the observation that the variant identified in our study as associated with increased RPVO is also associated with increased mRNA and protein expression of OSTN appears counterintuitive and warrants further investigation into the underlying functional mechanisms.

Using MR analyses leveraging plasma proteomic data, we observed that increased genetically predicted plasma levels of IL-1 R AcP protein, encoded by *IL1RAP*, were associated with reduced RVPO, highlighting IL1RAP-mediated pathway as a potential therapeutic target. IL1RAP is known to be a membrane co-receptor essential for the signaling of several pro-inflammatory cytokines, including IL-1 (α and β), IL-33, and IL-36 (83). Its soluble form can bind to these cytokines to resorb them, acting as a molecular “trap” that prevents them from attaching to pro-inflammatory cell receptors (83,84). Soluble sIL1RAP levels appear to promote the blocking of IL-1β, reducing its underlying production of granulocyte colony-stimulating factor which is associated with the development of thrombosis via the formation of neutrophil extracellular traps (85). As such, increased soluble IL1RAP levels would be directly linked to a decrease in RPVO, a result consistent with our MR findings. Several antibody-based strategies targeting IL1RAP, including nadunolimab and CAN10, have been developed in the context of inflammatory diseases and cancer to inhibit IL1RAP signaling (86). Our findings suggest that positive pharmacological modulation of IL1RAP activity could also represent a promising therapeutic avenue potentially serving as adjunctive therapy at initial phase of PE treatment to prevent the early development of RPVO. Sensitivity analyses further suggest that the two association signals observed at OSTN and IL1RAP represent statistically independent effects, despite both loci map to chr3q28. Beyond experimental studies aimed at elucidating the underlying biological mechanisms, long-read sequencing of the chr3q28 locus could help refine the genomic architecture of this region and clarify its relationship with RPVO. In addition, measuring IL-1 R AcP plasma levels in patients with unprovoked PE will be an important next step to clarify the potential value of this biomarker for predicting the risk of RVPO.

The chr3q27.3 region was detected by a haplotype analysis of two SNPs with marginal suggestive association (p<10^-3^) and mapping nearby the candidate *KNG1* locus. The identified risk-haplotype associated (p=2.96×10^-8^) with∼3-fold increase of RPVO covers a large genomic region including coding and non-coding RNAs among which several biological candidates can be identified such as *AHSG, HRG, KNG1* and *ST6GAL1*. *AHSG* has been notably reported to associate with plasma levels of coagulation factor V (87) and to have various vascular functions (88). *HRG* is known to be involved in rare forms of thrombophilia (OMIM #613116) but also to participate in thrombo-inflammation (89). By participating in the regulation of FXI, *KNG1* is also a strong candidate. However, plasma proteogenomic haplotype and MR analyses did not provide strong arguments in favor of one of these loci. Therefore, these analyses do not provide support for the use of pharmacological Factor XI inhibitors to prevent RPVO. Finally, by its role in regulating the inflammatory response and endothelial cell function (90,91), *ST6GAL1* is also a good candidate. All these elements clearly indicate the need for a more comprehensive genomic analysis of the chr3q27.3 locus to disentangle the detected association signal with RPVO.

Although not reaching our pre-specific genome-wide statistical threshold, we observed that the rare C allele of the *CCN4* rs16904846 was associated with a ∼4-fold increase of RPVO. This variant is a singleton with no LD (r^2^>0.50) with any nearby SNPs and was found associated with only CCN4 plasma protein levels. CCN4 codes for WISP1, an extracellular matrix protein that regulates cell proliferation, survival, and migration by modulating the extracellular environment. It is strongly involved in pulmonary fibrosis (74,75,92,93) and inflammation (94,95), promoting pathological tissue remodeling and inflammatory responses. Accumulating evidence suggests that WISP1 is involved in asthma and chronic obstructive pulmonary disease (COPD) (95), two clinical outcomes strongly associated with dyspnea, one of the main complications of RPVO.

Taken together, the findings at OSTN, IL1RAP and CCN4 provide strong genetic support to the key role of inflammation in RPVO, consistent with the inflammatory hypothesis raised by Shah et al (20). Additional support for this hypothesis comes from our MR analysis oof hematological traits, in which the strongest association was observed for eosinophil counts, whose role in inflammatory responses is well established (96), although this result did not survive correction for multiple testing. By contrast, our study provides little direct genetic support to the fibrinolysis hypothesis (17,32). Nevertheless, the suggestive association observed with *CPB2,* coding for TAFI, remains noteworthy. Future studies measuring plasma TAFI levels in unprovoked PE patients followed for RPVO would allow confirmation of the involvement of this fibrinolytic marker. In addition, IL1RAP-mediated regulation of IL-33 recently proposed as an inhibitor of tissue-factor pathway inhibitor (97), which itself has an emerging role in fibrinolysis (98), raises the possibility that IL1RAP could also be involved in RPVO mechanism through a potential inflammatory and fibrinolytic component.

We also observed suggestive associations at two platelets related genes (*GP5* and *GP1BA*), raising the possibility that platelet mediated pathways contribute to RPVO. These findings may warrant further investigation of anti-platelets strategies (99) as potential approach to prevent RPVO, particularly in genetically determined subgroups of patients or that the anti-platelets therapy in some PE patients may have influenced our findings. Unfortunately, we could not adjust our analyses for this information, since it was not documented in all our patients.

*CCN4* variant and IL1RAP PRS were also found to be associated with the risk of VTE recurrence, in a manner consistent with the patterns observed for RPVO. Then, these molecular findings provide encouraging insights into the mechanisms associated with RPVO and highlight potential therapeutic perspectives.

However, these overall results should be slightly tempered by several limitations. We here assembled the only three clinical cohorts of PE patients in which both RPVO and GWAS data were available to our knowledge, preventing formal replication in independent samples. Besides, heterogeneity across cohorts, including differences in mean age, time at which RPVO was measured, the proportion of PE patients with both DVT and PE (**Supplementary Table 4**), may have limited our chance to detect additional statistical findings. Furthermore, as we could not adjust our analysis for initial PVO measured at the time of acute PE, whose information was not available in all cohorts, we were not able to determine whether the observed genetic effects could exert early in the process of thrombus resolution. It is also important to note that detailed information on the nature of VTE recurrence—specifically whether it was provoked or unprovoked, and whether it presented as DVT or PE—was not available in the studies used. Another point worth mentioning is the very low incidence of CTEPH in our cohorts. This suggests that patients with RPVO and no pulmonary hypertension at early follow-up may represent a distinct population from those who will develop CTEPH over time. Accordingly, the study design may have limited our ability to identify molecular determinants of both RPVO and CTEPH. Finally, further work is needed to characterize the exact functional variant(s) at the identified loci and their downstream mechanisms that would lead to modulate RPVO.

In conclusion, this first GWAS on RPVO provides strong genetic evidence for the involvement of inflammation and vascular remodeling in RPVO and paves the way for future basic and translational studies in patients with unprovoked PE. Inflammation is a predominant mechanism and probably the key mechanism for RPVO, while impairment in fibrinolysis appears more marginal. Our results also highlight the potential ability to identify, among patients with RPVO, those who are at higher risk of recurrent PE involving important potential clinical implications.

## Acknowledgments

We warmly thanks Prof Chris Rhodes and Mark Toshner for facilitating access to the summary statistics of the GWAS for CTEPH (40). We are grateful to the Centre for Applied Genomics and The Hospital for Sick Children (Toronto, Canada) for their contributions to genotyping and data processing. Computational analyses of REVERSE I data were facilitated by the SciNET High Performance Computing Consortium at the University of Toronto and the Digital Research Alliance of Canada.

Statistical analyses of the EDITH, MARTHA, MEGA and PADIS-PE genomic data benefited from the technical support of the CBiB computing centre of the University of Bordeaux. In France, the EDITH MARTHA and PADIS-PE studies are part of the French Clinical Research Infrastructure Network on Venous Thrombo-Embolism (F-CRIN INNOVTE). In Canada participating centers are members of the CanVECTOR Network. These national networks are members of INVENT-VTE, the International Network of Venous Thromboembolism Clinical Research Networks (www.invent-VTE.com).

## Data availability

All data that support the findings in this study are available from the corresponding author upon reasonable request. GWAS summary statistics will be available in GWAS Catalog (GCST90860934).

## Authorship

Contribution: ADJ, D-AT, FC, FRR, GLG, NLS and P-EM participated in study concept and design. AvHV, CL, CR, FC, GLG, LG, MR, MT and SD participated in phenotype data acquisition or control quality. ADJ, AvHV, D-AT, FG, DB, GM, IC, J-FD, MG, NLS, OCLB, RO and SD, participated in genotype data acquisition or control quality. D-AT, FS, GM, KW, FG and OCLB participated in data analysis and interpretation. D-AT and FS wrote the initial draft of the manuscript which was reviewed and approved by all co-authors.

Correspondence: David-Alexandre Trégouët, 146 Léo Saignat – BPH U1219 Eleanor team – 33076 Bordeaux.

## Supplementary Figure legends

**Supplementary Figure 1. Distribution of RPVO values among REVERSE-I, EDITH and PADIS-PE studies.**

**Supplementary Figure 2. Quantile-Quantile plot of the GWAS meta-analysis results on RPVO (N=586).**

**Supplementary Figure 3. Region plot for the most strongly associated region in the GWAS meta-analysis on RPVO (N=586).**

