## Supplementary figures and images for "Genomic Evidence Links Inflammation to Residual Pulmonary Vascular Obstruction and Risk of Pulmonary Embolism Recurrence"

### Supplemental Figures

#### Supplementary Figure 1

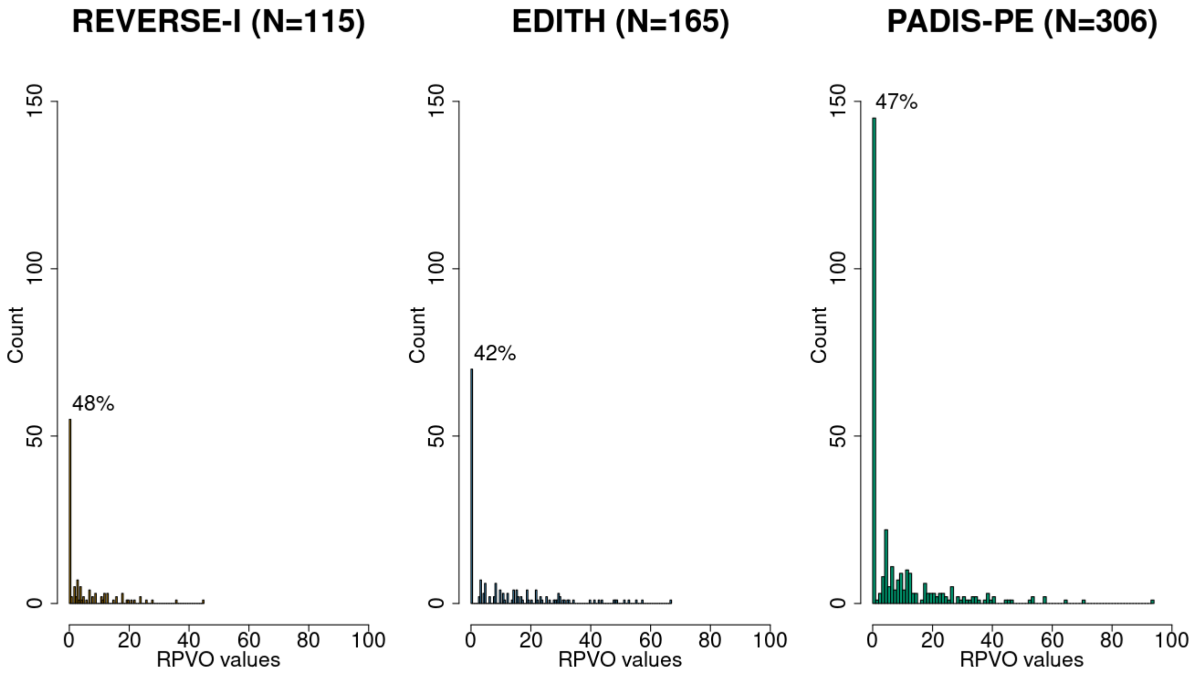

##### Supplementary Figure 2

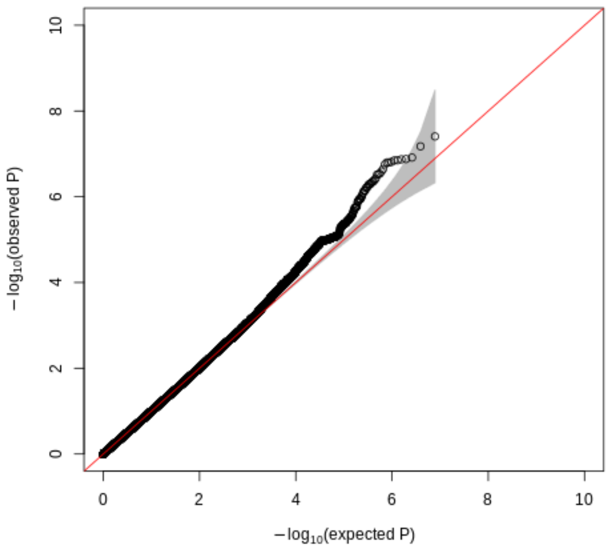

#### Supplementary Figure 3

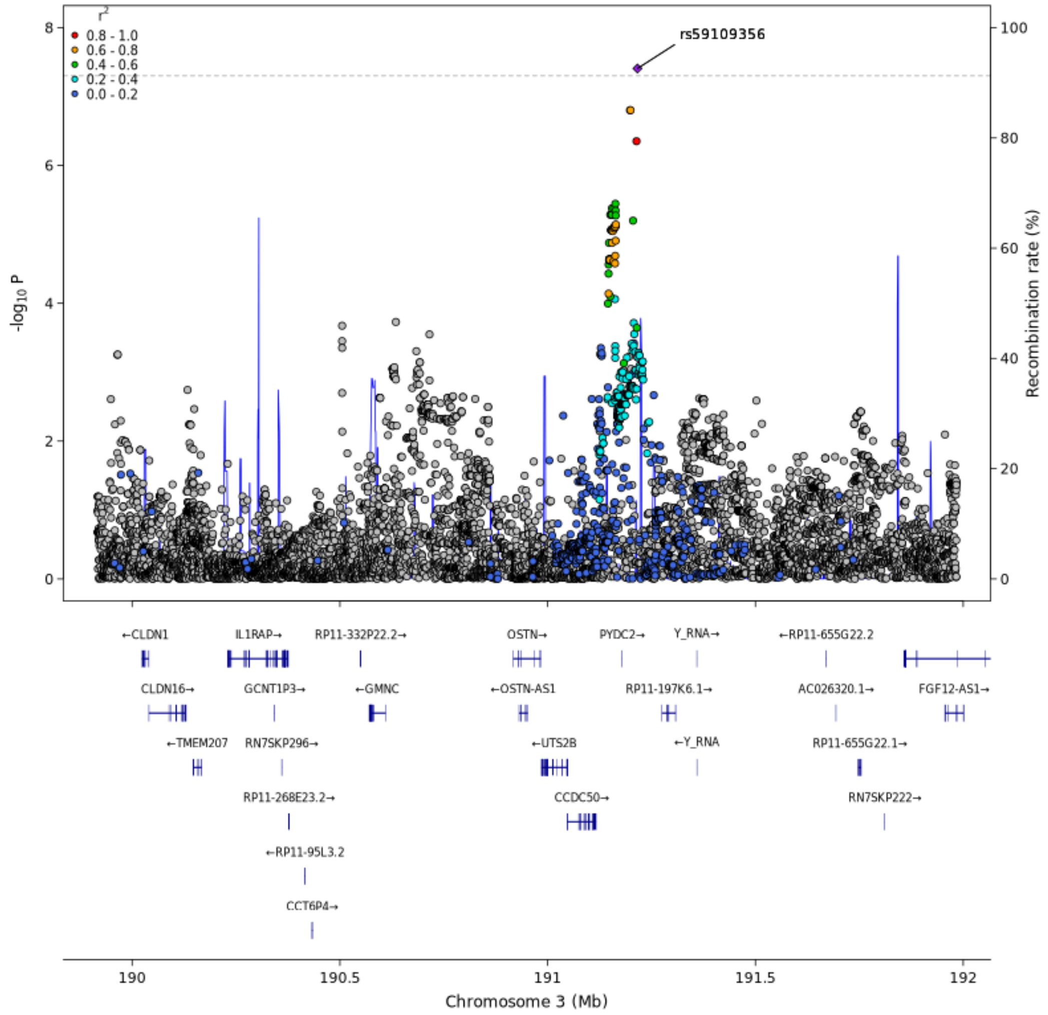
